# Heart rate variability as biomarker for bipolar disorder

**DOI:** 10.1101/2022.02.14.22269413

**Authors:** Andrea Stautland, Petter Jakobsen, Ole Bernt Fasmer, Berge Osnes, Jim Torresen, Tine Nordgreen, Ketil J Oedegaard

## Abstract

Bipolar disorder (BD) is characterized by alterations in mood, energy levels and the ability to function. Accordingly, it is also associated with dysfunction of the autonomic nervous system (ANS), indexed by heart rate variability (HRV). Literature concerning differences in ANS functioning between mood states is still sparse. The main aim of the study was to investigate within-individual changes in HRV from manic to euthymic states in bipolar disorder (BD). This is the first study to do so using wrist-worn sensors. Seventeen patients with BD were equipped with photoplethysmography (PPG) sensor wristbands and provided 24-hour recordings both during a manic state and a euthymic state. We calculated mean heart rate and the commonly used HRV measures SDNN, RMSSD, HF, LF and Sample Entropy in 5-minute segments during rest at night. We compared HRV by mood state within individuals using paired t-tests and linear regression to control for age and sex. Recordings from 15 BD patients were analyzed. There were statistically significant increases in HRV measures SDNN, RMSSD, LF and Sample Entropy from mania to euthymia. Effect sizes were predominately large. Our findings reveal lower HRV in the manic state compared to the euthymic state. This indicates that HRV collected by wrist-worn PPG sensors is a possible biomarker for bipolar mood states. Movement artifacts were problematic and sampling during rest or in combination with actigraphy is recommended. Our findings can be further implemented to develop a monitoring device for bipolar patients.

## Introduction

Bipolar disorder (BD) is a severe mental disorder associated with intense mood fluctuations, lifelong disability and increased mortality. It affects about 1-3% of the population, with a lifetime suicide prevalence of 15-20% [1]. Patients typically need long term follow-up, mood stabilizing medications and psychoeducation to reduce the frequency of new mood episodes and achieve mood stability [2, 3]. Clinical assessments of BD symptoms are limited to subjective evaluations by clinicians and patient recall bias and poor illness insight [4]. Consequently, objective methods for monitoring state changes are sought-after, and new tools to improve early detection and intervention in BD are in demand [5].

The autonomic nervous system (ANS) controls a multitude of bodily functions and links the central nervous system to peripheral organs such as the heart [6-8]. Heart rate variability (HRV) reflects fluctuations in time intervals between consecutive heartbeats and is a well-documented and studied measure of ANS activity. Studying HRV allows us to observe the adaptive abilities of the heart and ANS to environmental changes [9]. HRV measures are commonly calculated using time-domain, frequency-domain or non-linear approaches. Through these methods, it is possible to observe the balance between the “fight-or-flight” sympathetic and “rest-and-digest” parasympathetic branches of the ANS [10-12].

The adaptiveness of the ANS is well-studied within psychology and psychopathology. High HRV scores have been found correlated with positive traits such as sustained attention and communication abilities. Reduced vagally mediated HRV has been found across several different psychopathological conditions and is therefore suggested to be a possible transdiagnostic biomarker for mental illness [13].

BD is characterized by changes in activation, and correspondingly, ANS activity and function has been found to be altered and dysregulated in BD [10, 14-19]. A 2017 review article and meta-analysis reported reduced HRV in BD compared to healthy controls [10]. Moreover, two recent studies report an inverse relationship between BD illness severity and HRV [19, 20]. This is of interest considering diagnostic tool development, general understanding of the disorder, and as a candidate biomarker of cardiovascular risk [21].

HRV differences between affective states have been sparsely studied, and review articles reveal that most of the literature investigates group level differences [10, 17]. Only one prior study has investigated within-individual HRV changes related to mood states. This 2018 study compared 19 manic males to their euthymic selves, finding HRV changes suggestive of ANS improvement [22]. A small observational study found increased HRV in mania compared to euthymia and depression on a group level, opposing most of existing literature [23]. Finally, a group has investigated HRV changes in the transition from pathological to euthymic states in ten bipolar patients, finding a steady increase as symptoms subsided [24]. The same group used innovative technology to accurately classify affective states in long-term HRV analysis of eight BD patients [25]. These findings indicate that HRV can be used as an objective biomarker for distinguishing between affective states and monitoring the course of bipolar illness. However, more studies on state dependent HRV changes within individuals as are needed. Based on this existing literature, we hypothesized lower HRV in mania compared to euthymia.

### Aims of the study

To identify within-individual differences in heart rate variability measures between manic and euthymic states in bipolar patients. We used high-quality wrist-worn pulse sensors in a natural patient setting to evaluate the applicability of such a device in bipolar patient monitoring.

## Materials and methods

### Design

This HRV study is part of an ongoing clinical study consisting of two parts: an intra-individual inpatient case control study, and a prospective outpatient observational study.

### Sample

Eligible probands were individuals with BD residing in the catchment area of Haukeland University Hospital in Bergen, Norway. The inpatient part of the study required an ongoing manic episode and admission to Haukeland University Hospital. Outpatients for the prospective observational study were included from September 2018 up to and including November 2020. Inpatients were included from November 2017 up to and including May 2020.

Inclusion criteria were Norwegian speaking individuals 18 to 70 years old diagnosed with BD by the ICD-10 criteria, able to comply with instructions and cognitive abilities clinically estimated to correspond to an IQ above 70. All participant diagnoses were set or confirmed by resident physicians or senior consultant psychiatrists at Haukeland University Hospital. Exclusion criteria were participation refusal, previous head trauma requiring hospital treatment, an organic brain disorder, ongoing substance dependence (nicotine permitted) or being in a state of withdrawal.

### Procedure

Inpatients were invited to participate upon referral from the psychiatry residents of the two closed affective wards at Haukeland University Hospital. Inpatient participants were informed about and invited to continue into the outpatient study. We also recruited participants for the outpatient study from the hospital outpatient clinic, through the local advocacy group for BD, and psychoeducational courses hosted by the hospital. No financial compensation or treatment perks were provided.

We assessed the included patients and obtained 24-hour wristband sensor recordings at the time of inclusion. Inpatients were reassessed and recorded at discharge from closed or at the open ward they were transferred to, when in remission. Outpatients were followed routinely for one year with mood assessments. Changes in mood state triggered 24-hour wristband sensor recordings.

### Measures

#### Heart rate variability

Empatica E4 devices with photoplethysmography (PPG) sensors have been validated against ECG for HRV purposes and were used for pulse detection. Heart rate was monitored for 24 hours in a naturalistic setting at a 64Hz sampling rate [26, 27]. Research personnel secured the device on the participants’ dominant wrist as snugly as tolerated, ideally to a fit allowing one finger under the band. The dominant hand was recommended to us by the manufacturer. The participants’ only instructions were not to shower or touch the device during the recording. We used Empatica’s online software, E4 Connect, to visualize gross loss of sensor contact, as identified by flattened skin conductance and temperature measurements and peaks in heart rate, and to download the inter-beat-intervals (IBI) data as comma-separated value files for post-processing.

IBI data was analyzed in Kubios HRV Premium software version 3.4.2 [28]. We set the artifact correction threshold to automatic and applied the smoothing priors detrending method (*λ*=500) [28]. We located 5-minute samples of high quality manually using a systematic approach. As artifact rates in PPG recordings are lowest during rest, we focused on night-time samples during sleep, cross-checking the accelerometer data from the device as assurance of rest [27]. We set the artifact correction cut-off to the recommended 5% and considered data with a higher correction rate as poor quality sections. All analyses of HRV were performed on samples from as close to initiation of sleep as possible, ranging from 8.10 p.m. to 5.10 a.m. (mean 11.34 p.m., standard deviation 112 minutes.)

We selected commonly used HRV measures from all three domains. From the *time domain* measures, we selected root mean square of the successive differences (RMSSD), standard deviation of average R-R intervals (SDNN) and mean heart rate (HR). We viewed RMSSD as our main HRV measure as it mirrors the variance in time between heartbeats, is widely used for monitoring vagally mediated (i.e. parasympathetic) HRV changes, and is well-suited for use on our 5-minute data segments [29]. In the *frequency domain*, we chose to examine high frequency (HF) and low frequency (LF) oscillations (ms^2^/Hz). HF is perceived as reflective of predominately parasympathetic activity, while LF interpretation remains debated and involved in both parasympathetic and sympathetic activity [30]. The frequency measures were presented in logarithmic form of results from an autoregressive approach. We selected Sample Entropy (SampEn) as the primary measure from the *non-linear domain*. It reflects the complexity and degree of chaos in the heart rate series [29].

#### Clinical assessment

Mood was evaluated using the Young Mania Rating Scale (YMRS) [31, 32] and Montgomery Asberg Depression Rating Scale (MADRS) [33, 34], two commonly used evaluation scales for mania and depression in clinical and research settings. We defined euthymia as a total YMRS score < 10 [32, 35]. Diagnosis was confirmed by research personnel trained in the use of Mini-International Neuropsychiatric Interview (M.I.N.I.) [36], when the subjects were euthymic.

### Statistics

We used paired two-tailed t-tests to explore the significance of mood state on mean HR and HRV features of interest: RMSSD, SDNN, LF, HF and SampEn. Significance levels were set to p = 0.008, by the Bonferroni method (*α* = 0.05), due to the small number of subjects (N = 15) relative to the number of tests (six). This was done to control for false positive findings due to multiple testing. Prior to analysis, RMSSD values were log transformed due to distribution skewness. Effect sizes of change by mood state were calculated using Hedges’ g. Sex and age are customary confounders of HRV [29]. We ran separate linear regression models for each of our five HRV measures to examine confounding effects. The dependent variables of the linear regression models were the manic-euthymic differences of the HRV measures (e.g. RMSSD) by turn, and the independent variables were sex and age. Due to the small number of subjects, we did not include mood state as an independent variable in the linear regression models, using the models purely to examine confounding effects.

### Ethics

The Norwegian Regional Medical Research Ethics Committee West approved the study (2017/937).

## Results

### Background characteristics

We invited 51 inpatients to participate, of which 34 accepted. 17 of the 34 completed both assessment points (manic and euthymic). We included 35 participants to the observational part of the study. Two of the 35 provided recordings from both a euthymic and manic state. Ultimately, 15 participants provided recordings that passed quality control and were included for analysis. See Table 1 for a description of the study participants and clinical mood states.

**Table 1.**
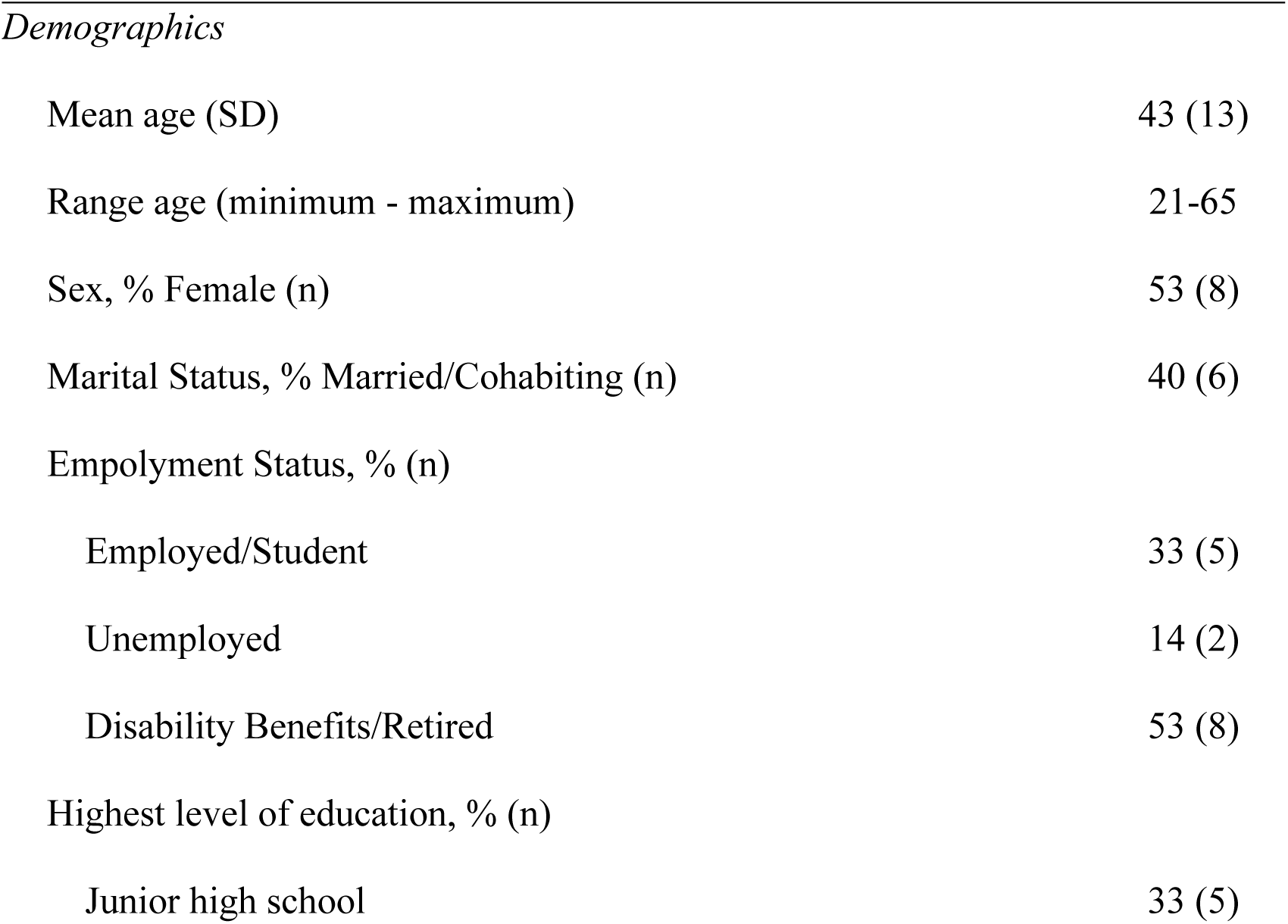
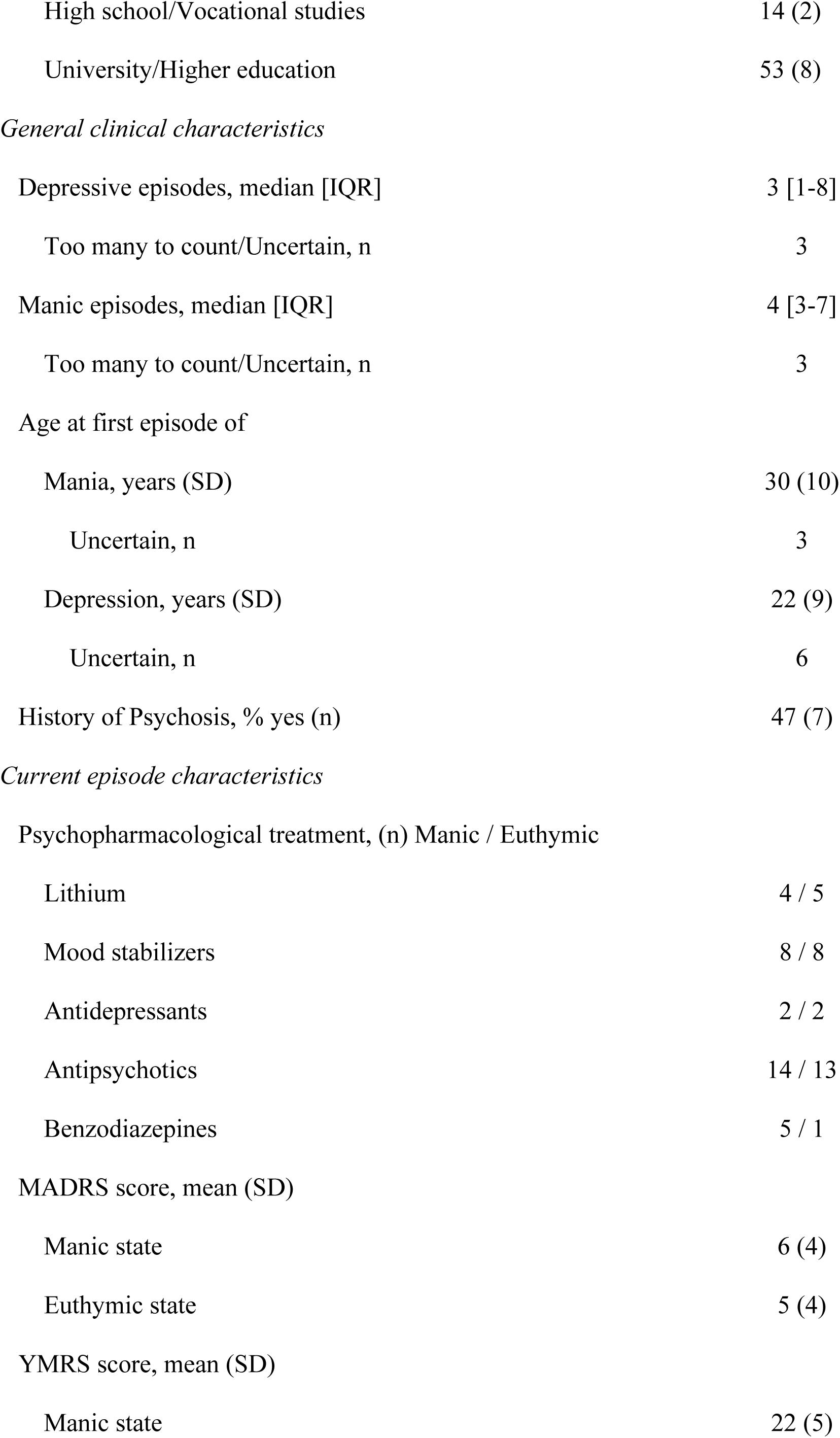
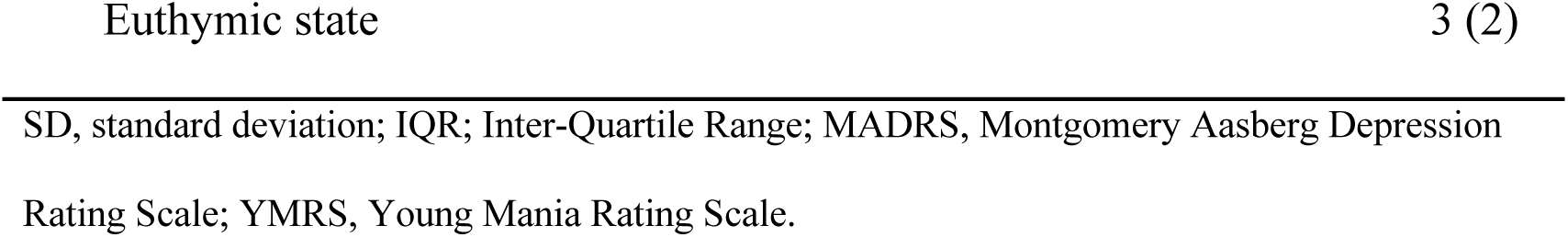
Characteristics of N = 15 bipolar type 1 study participants at baseline and clinical characteristics during a manic and euthymic state.

### Analysis results

Considerable changes in heart rate and several HRV measures were observed from mania to euthymia, with overall large effect sizes (Fig 1). We found a statistically significant decrease in HR with a large effect size (g = 0.92), although the range was large in the manic state (49-102 beats per minute). Time domain HRV measures SDNN and RMSSD increased significantly (g = 1.01; g = 1.13) from manic to euthymic state, as did the frequency measure LF (g = 1.24) and the non-linear measure Sample Entropy (g = 0.80). HF also increased from mania to euthymia, but the change was not statistically significant.

**Fig 1.**
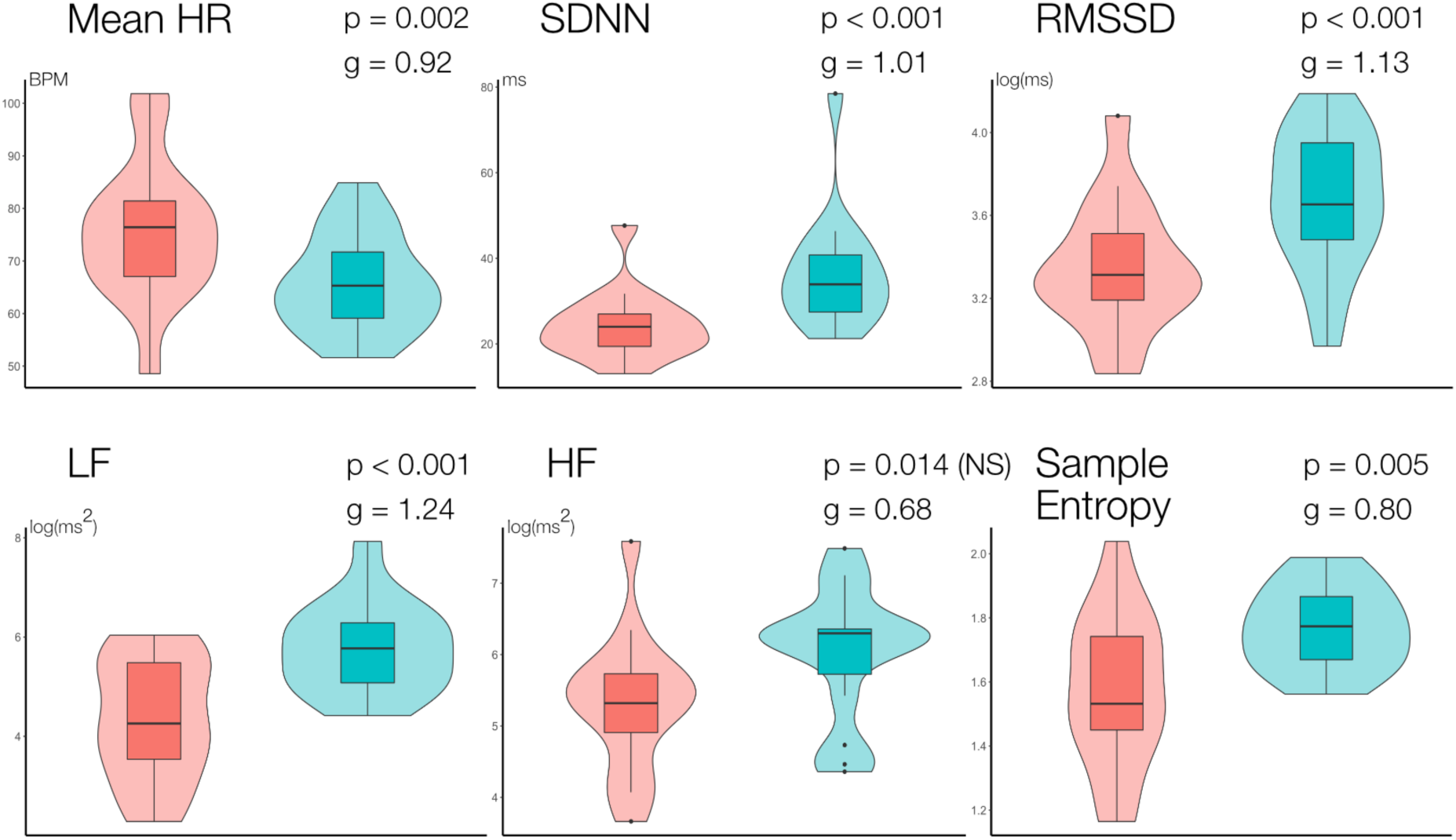
Paired comparison of HRV measures by mood state in 15 bipolar subjects. Paired t-tests with p = 0.008 significance levels. Violin plots show sample distributions while boxplots show median and variance. NS = not statistically significant.

The linear regression models revealed a possible statistical effect of age on SDNN (estimate = -0.67 per added year, p = 0.02). No confounding effects were identified for the remaining measures (see S1 Table).

## Discussion

There is a lack of disease monitoring options in bipolar disorder, and HRV is a promising biomarker candidate. This study found a decrease in several HRV measures and increased heart rate during mania when compared within individuals to their euthymic selves. It is one of few studies examining state related changes longitudinally within bipolar subjects, and the first to employ a user-friendly wrist worn device. These features are significant when considering innovation possibilities. Our results are suggestive of decreased vagally mediated parasympathetic activity during bipolar mania.

HRV differences were observed in measures from the time, frequency and non-linear domains. From manic to euthymic state SDNN, RMSSD, LF and SampEn all increased significantly with predominately large effect sizes. This is largely in agreement with a recent study with a design and sample size resembling ours, although the only shared measurements of statistical significance between our studies are RMSSD and SampEn [23]. Their study approach was more controlled than our natural design, and there were several methodological differences between our studies. First, they used an ECG chest strap for heart rate measurement, instructing the participants to lie still for 20 minutes. Our study used a lightweight wrist-worn PPG device and a natural approach, examining recordings made during sleep. Second, they excluded patients with high motor activity and all participants were men. This could result in slightly different clinical profiles and a bias toward higher HRV scores due to the male sex [37]. They also restricted medication intake to aripiprazole and lithium, while we did not interfere with treatment. They did not control for possible confounders. The similarity of our results, despite some methodological differences, emphasize the robustness and biological underpinning of our shared findings.

Larger group-level studies have also reported decreased HRV variance and entropy measures in manic patients when compared to healthy controls [16, 38]. They used ECG-based measuring methods. Henry et al. ultimately posed the question of whether low HRV is due to a bipolar trait or also the manic state. Our findings confirm their theory of low HRV being a consequence of the manic mood state, not simply the underlying illness [38].

Lastly, we found a decline in heart rate in euthymia compared to mania (g = 0.82). This has been described in previous literature [16, 23, 38]. Heart rate is regulated by several mechanisms which are altered during a manic episode. Emotional and physiological stress promote sympathetic upregulation in the limbic system, and secretion of stress hormones [39]. Higher HR during mania is therefore not a surprising discovery. One could argue that the higher HR during mania could be due to increased motor activity, and consequentially higher HRV scores. However, we did not discover a significant difference in the amount of movement, as measured by actigraphy in this sample [40].

During the course of our study, methodological weaknesses became apparent. First, the representability of our participants should be addressed. The design requires an active role from treating physicians to refer manic inpatients capable of cooperating. This excludes subtypes of BD with paranoid traits or rapid mood changes, and those overly aggressive or cognitively affected. Substance abuse is also a common comorbidity in BD, here excluded. Due to the nature of scientific work, with ethical requirements and attempts at biological classification, we believe that such a narrowing of scope is necessary at this stage. This is also the case for comparable studies, as a certain degree of cooperation is required, and comorbidities are commonly excluded to provide a clearer image of effects attributed solely to BD.

Still, our sample contained older participants (43 ±13 years) compared to similar studies and they were more educated than expected [41].We believe that this is partly due to our recruitment method, appealing more to their inherent interest in their illness and contribution to society. We experienced a higher degree of drop-out in younger participants as their manic symptoms subsided. Such patients could for instance be lost due to a sudden repeal of coerced treatment and subsequent ceased contact. A possible explanation is that both more mature and educated BD patients are more likely to be interested in contributing to increased knowledge and grounded enough to complete the study.

The effect of psychiatric drugs on HRV is controversial and study findings vary largely due to heterogeneous methodology. Overall, the effect of psychiatric drugs on HRV has been considered miniscule compared to the underlying HRV disturbances in psychiatric disorders. The exceptions are tricyclic antidepressants and the atypical antipsychotic clozapine [38, 42, 43]. The patients in our sample were roughly on the same medication classes during manic and euthymic recordings, see Table 1. Although dosages are not considered, this likely implies smaller effects of medication differences on HRV by mood state. In addition to the similarities between our findings and those of the previously mentioned study restricting medication use, similar results have been described during unmedicated mania, further solidifying our results [16, 23]. As the participants are considered their own controls in this natural study and the literature is not adamant about controlling for medication use; we did not stress the effects of antidepressant and antipsychotic medications on HRV.

Benzodiazepines are nonetheless an exception, as its use is more widespread during mania than euthymia. The effects of benzodiazepines on HRV are, however, sparsely studied. HRV has been found to drop shortly after IV administration of benzodiazepines [44]. Contrarily, regular use in older individuals did not alter HRV [45]. Although we intuitively believe that benzodiazepines could affect HRV, we did not have the statistical power needed to perform a reliable analysis on such an effect. The conflicting literature and the large effect sizes in our results supported not pursuing this issue further. Research into the effects of benzodiazepines on HRV in both BD patients and patient controls is plainly lacking and could elucidate benzodiazepine mechanisms of action while also improving HRV study accuracy in patient samples.

Known weaknesses of PPG-devices such as the Empatica E4 are susceptibility to artifacts during movement and cognitive and emotional stress [27]. Despite proper fitting and instructions, we found that a selection of the 24-hour recordings were not of satisfactory quality for HRV analyses. This was especially true for manic recordings. However, we discovered a pattern of overall higher quality during rest at night. In order to keep as many participants as possible in the analyses, we therefore focused on night recordings. This choice provided a new caveat – the effect of circadian rhythms on HRV. In healthy individuals HRV varies throughout the day and reaches a high-point around 03.00 a.m. [46]. In theory, due to our participants falling asleep at different times (mean 11.34 p.m., standard deviation 112 minutes) both across mood states and individuals, our recordings could be subject to circadian bias. Still, given the established circadian disruptions associated with BD, it is questionable if standardizing the measurement time would insure similar conditions between subjects.

### Strengths of the Study

The major strength of our study is the use of a within-individual design, wherein each participant functions as their own control. Within-individual designs provide a higher degree of statistical power than groupwise comparisons. This is also the only study of its kind using PPG devices. Finally, this study design looks at real-life values not recorded under strict instructions. In the context of disease monitoring at home, real-life data is crucial.

## Conclusion

We found a large increase in several heart rate variability measures and a decrease in heart rate from a bipolar manic to euthymic state. This is the first study to describe such within-individual HRV changes utilizing a wrist-worn PPG device. The effect sizes of our results were large and are in line with the majority of previous literature on the subject. Consequently, HRV could act as a biomarker for monitoring bipolar shifts in mood state. We believe that wrist-worn heart rate monitoring holds innovation potential for monitoring bipolar disorder.

### Future work

Future work should consider coupling PPG and actigraphy monitoring with an algorithm to automatically measure heart rate and calculate HRV when the subject is in a resting position, monitoring throughout the day. Moreover, focusing on non-hospital samples not yet in treatment and the transitions between mood states would be of value, as a monitoring device would be most efficient if able to detect *shifts* in mood. Finally, multi-sensor machine learning approaches show promise. We are currently collaborating with the ROBIN group at the University of Oslo on mood-state recognition based on multi-modal sensor data [47].

## Data Availability

A minimal dataset is included in the supplementary files. Is is unidentified following ethical standards.

## Acknowledgments

We would like to thank statistician Christopher Andreas Bartz-Johannessen for statistical consultation. This publication is part of the INTROducing Mental health through Adaptive Technology (INTROMAT) project.

## Supporting information

**S1 Dataset. Output from HRV analyses**. Data contains analysis results of all HRV variables investigated from one manic and one euthymic sample from each participant (N = 15). De-identified and restricted due to ethical and patient privacy concerns.

**S2 Table. Results from linear regression analyses**. Investigating confounding effects of age and sex on heart rate and heart rate variability variables. The dependent variables of the six linear regression models were the manic-euthymic differences of the HRV measures, and the independent variables were sex and age. Standard error shown in parentheses.

