## Supplementary material for "Heart rate variability as biomarker for bipolar disorder": S2: Supplemental Table

**S2 Table. Results from linear regression analyses.**

Investigating confounding effects of age and sex on heart rate and heart rate variability variables. The dependent variables of the six linear regression models were the manic-euthymic differences of the HRV measures, and the independent variables were sex and age. Standard error shown in parentheses.

| | Mean HR | SDNN | RMSSD | LF power | HF power | Sample entropy |
|---|---|---|---|---|---|---|
| Age, mean centered | -0.28 | -0.67 * | -0.01 | -0.01 | -0.02 | 0.00 |
| | (0.24) | (0.24) | (0.01) | (0.03) | (0.03) | (0.01) |
| Sex, male | -2.81 | 11.46 | 0.06 | 0.36 | -0.06 | -0.09 |
| | (5.85) | (5.85) | (0.17) | (0.70) | (0.61) | (0.14) |
| nobs | 15 | 15 | 15 | 15 | 15 | 15 |
| r.squared | 0.19 | 0.40 | 0.08 | 0.02 | 0.08 | 0.03 |
| adj.r.squared | 0.06 | 0.30 | -0.07 | -0.14 | -0.08 | -0.13 |
| sigma | 9.69 | 9.69 | 0.28 | 1.17 | 1.01 | 0.24 |
| statistic | 1.45 | 4.00 | 0.54 | 0.15 | 0.49 | 0.21 |
| p.value | 0.27 | 0.05 | 0.60 | 0.87 | 0.63 | 0.81 |
| df | 2.00 | 2.00 | 2.00 | 2.00 | 2.00 | 2.00 |
| df.residual | 12.00 | 12.00 | 12.00 | 12.00 | 12.00 | 12.00 |
| nobs.1 | 15.00 | 15.00 | 15.00 | 15.00 | 15.00 | 15.00 |

*** $p < 0.001$; ** $p < 0.01$; * $p < 0.05$.
